# Pre- and Post-Portosystemic Shunt Placement Metabolomics Reveal Molecular Signatures for the Development of Hepatic Encephalopathy

**DOI:** 10.1101/2023.01.02.22281374

**Authors:** Ana Carolina Dantas Machado, Stephany Flores Ramos, Julia M. Gauglitz, Anne-Marie Carpenter, Daniel Petras, Alexander A. Aksenov, Un Bi Kim, Michael Lazarowicz, Abbey Barnard Giustini, Hamed Aryafar, Irine Vodkin, Curtis Warren, Pieter C. Dorrestein, Ali Zarrinpar, Amir Zarrinpar

**Author notes:** Co-Corresponding author: Ali Zarrinpar, M.D., Ph.D., Amir Zarrinpar, M.D., Ph.D. Contributed equally (Ana Carolina Dantas Machado, Stephany Flores Ramos).

## Abstract

Hepatic encephalopathy (HE) is a common complication of advanced liver disease causing brain dysfunction. This is likely due to the accumulation of unfiltered toxins within the bloodstream. A known risk factor for developing or worsening HE is the placement of a transjugular intrahepatic portosystemic shunt (TIPS), which connects the pre-hepatic and post-hepatic circulation allowing some blood to bypass the dysfunctional liver and decreases portal hypertension. To better understand the pathophysiology of post-TIPS HE, we conducted a multi-center prospective cohort study employing metabolomic analyses on hepatic vein and peripheral vein blood samples from participants with cirrhosis undergoing elective TIPS placement, measuring chemical modifications and changes in concentrations of metabolites resulting from TIPS placement. In doing so, we identified numerous alterations in metabolites, including bile acids, glycerophosphocholines, and bilirubins possibly implicated in the development and severity of HE.

## INTRODUCTION

Hepatic encephalopathy (HE) is a common complication of advanced liver disease, with up to 30-40% of patients developing this neurological condition as their disease progresses.^1^ The phenotype of HE can vary widely from mild cognitive impairment to more serious symptoms such as disorientation and coma. While the etiology of HE is complex, the most prevalent theory of its pathophysiology is the inability of dysfunctional hepatocytes to process enteric neurotoxins that accumulate within the splanchnic circulation, allowing them to enter the systemic circulation, and precipitate cerebral inflammation and cerebral edema. ^2,3^

Due to the elevated pressure within the portal venous system and resultant hepatofugal flow, extrahepatic collateral blood vessels known as varices form to allow for portal blood return to the vena cavae and the systemic circulation.^4^ Porto-hepatic venous collaterals, or intrahepatic shunts, can also form, although efforts to characterize this phenomenon have been sparse.^5–7^

In severe liver disease with refractory ascites or variceal bleeding, transjugular intrahepatic portosystemic shunt (TIPS) placement is a therapeutic intervention that can reduce pathologies caused by portal hypertension. With TIPS, a synthetic shunt connects the portal venous system to the hepatic venous system, such that some blood can bypass the cirrhotic liver and reduce portal hypertension and its sequelae. Although TIPS can significantly improve the morbidity and mortality of patients with cirrhosis and increase transplant-free survival, TIPS placement is also a known risk factor for the development of HE and, thus, preexisting HE is a relative contraindication to TIPS placement. TIPS can increase the incidence of HE by 30-55% post-procedure, with 90% of affected patients developing it within the first three months.^8–11^

While the association between TIPS placement and HE exacerbation is well-documented, the mechanism is poorly understood. Ammonia level is usually associated with HE, but studies examining ammonia level and HE development are conflicting. The association is highly dependent on the chronicity of hyperammonemia and varies greatly in outcome and thus levels are not typically followed clinically.^12–15^ Therefore, neurotoxic compounds beyond ammonia likely contribute to HE. Microbial products such as aromatic amino acids and bile acids (BAs) are candidate neurotoxins, but it is still unclear which microbial products or metabolites induce HE onset and how TIPS placement affects the concentration of these compounds within the systemic circulation.^3,16–18^

A promising method to identify such metabolites is untargeted metabolomics, which has been extensively applied to study microbial community function and host-microbe interactions.^19– 24^ Targeted metabolomics allows for absolute quantification of specific metabolites such as amino acids and fatty acids and is limited to a few previously identified compounds for which authentic standards are available. Untargeted metabolomics, on the other hand, allows for measurement of both known and unknown metabolites, thus enabling discovery of novel metabolites and insights into biological function.^21,25^ A key recent advance in untargeted metabolomics is the development of ion identity molecular networking (IIMN) to expand the annotation and quantification of metabolite features by enabling feature correlation analysis coupled with matching among spectral libraries and molecular networking.^26–31^ Even for features that cannot be fully annotated, molecular networking can be used to infer compound subclass level annotation (e.g. bilirubin, bile acid) through the network connections. The potential breadth of information that can therefore be gained from untargeted metabolomics with these recently developed bioinformatic tools can lead to the identification of novel therapeutics and biomarkers.^22,32–36^ Moreover, by obtaining serial samples from a single subject, investigators can prioritize proportional changes between biosynthetically related compounds, and postulate putative biotransformations, and what effects these changes may have on overall physiology.^37^

In this study, we performed an untargeted metabolomic analysis on hepatic vein and peripheral vein blood samples from patients undergoing TIPS. Our aim was to understand which metabolite levels change as a result of portosystemic shunt placement and whether these changes can predict worsening HE. To do so, we used standard and innovative untargeted metabolomics analyses that increased the overall set of annotations and allowed an exploration of the potential chemical modifications undergone by, and changes in levels of, the compounds after portosystemic shunt placement. We hypothesize that early shifts in these compounds predict the development and severity of HE and suggest the presence of intrahepatic shunting in some cirrhotic livers that is predictive of future development of HE.

## RESULTS

### Characterization of metabolites before and after TIPS placement implicates plasma bile acids levels

A total of 22 participants underwent TIPS placement at one of the two centers (**Supplementary Fig. 1**). Just prior to TIPS placement, 20 participants provided a peripheral vein (PIV) sample (**Fig. 1a**). During the TIPS procedure, the interventional radiologist collected hepatic vein (HV) blood for 20 participants just prior to and just after shunt placement. After the procedure but prior to admission to the post-anesthesia care unit, 20 of the participants provided another PIV blood sample. Finally, 20 of the participants provided a fasting PIV blood sample on the day of discharge with their morning labs (i.e., before discharge, BDc) (**Fig. 1a**). In total, 19 individuals had both pre- and post PIV, and 18 individuals had pre- and post-PIV and HV samples, thus allowing paired analysis for many of our comparisons. Participants were monitored by chart review for up to a year after their procedure and the worst HE grade during that period for each participant was noted. For two of the individuals who were readmitted with HE exacerbation during the study period, additional PIV blood samples were also obtained. An overview of participant demographics and clinical characteristics is described in **Supplementary Table 1**.

**Fig. 1:**
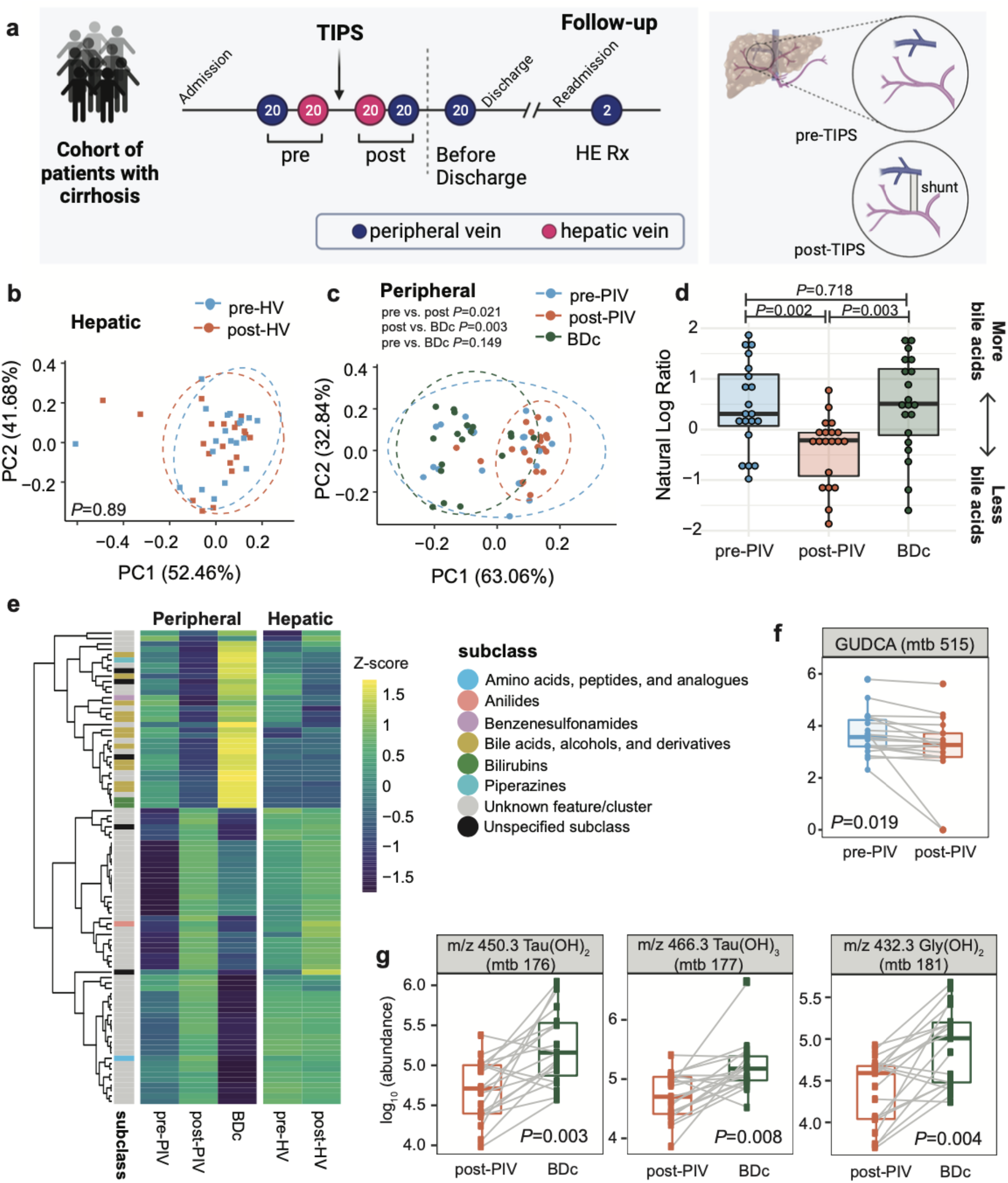
The effect of TIPS placement on the hepatic and peripheral metabolomes. **a**, Blood sample collection design. PIV (purple) and HV (pink) blood samples were collected pre- and post-TIPS, and following readmission for HE treatment. Visual of the location a shunt is placed during TIPS. Robust principal component analysis (RPCA) showing metabolome dissimilarities across participants in (**b**) pre-HV and post-HV samples and (**c**) pre-PIV, post-PIV, and BDc samples. **d**, The natural log ratio of bile acids to bilirubins. **e**, Heatmap showing the 87 unique metabolites that were significantly different between at least two of the three timepoints (paired Wilcoxon, FDR corrected at q-value 0.05); 25 unique metabolites significantly different pre vs. post-TIPS and 84 significantly different post-PIV vs. BDc (FDR < 0.1). **f**, Pre- vs. post-PIV levels of GUDCA, (paired Wilcoxon *P*=0.019). **g**, Examples of bile acids that were significantly different post-PIV vs. BDc (paired Wilcoxon, FDR corrected at q-value 0.05). (GUDCA=Glycoursodeoxycholic acid; m/z 450.3 Tau(OH)_2_ (mtb 176), m/z 466.3 Tau(OH)_3_ (mtb 177), and m/z 432.3 Gly(OH)_2_ (mtb 181) are annotated as bile acids through suspect library matching).

We interrogated differences in molecular distributions revealed by untargeted metabolomic associated with TIPS and HE severity. To do so, we characterized the metabolome of blood plasma samples collected directly from hepatic and peripheral veins pre- and post-TIPS (pre-PIV, pre-HV, post-HV, post-PIV), as well as before discharge (BDc). Metabolite features identified through untargeted LC/MS-MS were clustered by MS/MS similarity and annotated with a spectral library using feature-based molecular networking (FBMN) (**Supplementary Fig. 2a**) through the Global Natural Products Social Molecular Networking (GNPS) platform (https://gnps.ucsd.edu/).^27^ This resulted in the annotation of 234 features (39.3%), referred to as metabolite, among several clusters within the network (**Supplementary Fig. 2b-c**). This annotation rate which greatly exceeds typical annotations (below 10%) is enabled through the use of Nearest Neighbor Suspect Spectral Library.^38^

Using the untargeted metabolomic data, we investigated how TIPS placement affects the metabolome in the peripheral and hepatic vein samples (**Fig. 1b-d**). Metabolome dissimilarities across participants (as measured using the robust Aitchison β-diversity) pre- to post-HV were not significantly different (PERMANOVA *P*=0.89, **Fig. 1b**), but the difference reached significance between pre- and post-PIV samples (PERMANOVA *P*=0.021, **Fig. 1c**). To account for post-TIPS changes that may take longer to manifest, we assessed dissimilarity of the BDc samples to our earlier PIV samples and observed no change pre-PIV to BDc (PERMANOVA P=0.149) but a strong change post-PIV to BDc (PERMANOVA *P*=0.003, **Fig. 1c**). These results indicate that the biggest metabolomic changes in our samples did not occur immediately after TIPS placement but rather after TIPS placement and before discharge.

Since the most significant metabolome changes occured between the post-PIV to BDc metabolome, we interrogated which metabolites are driving these shifts. To do so, we used Qurro to visualize the log-fold change (rank) of metabolites contributing to this difference.^39^ This analysis demonstrated that bile acids as a group were major contributors to the dissimilarity (**Supplementary Fig. 3**). To better quantify this shift in bile acids, we used the natural log ratio of bile acids to bilirubins, a group of metabolites similarly present in all three PIV timepoints (**Supplementary Fig. 3**). This analysis demonstrated that the abundance of bile acids drops immediately after shunt placement, but abundances are restored to pre-PIV level by the time of discharge (**Fig. 1d, Supplementary Fig. 3**).

Bile acid changes across different time points along the TIPS procedure are also shown in paired Wilcoxon analysis of individual metabolites. Twenty-four metabolites were significantly different pre- vs. post-PIV while 12 were different pre- vs. post-HV. (FDR <0.1; **Fig. 1e, Supplementary Table 2**). These two sets of metabolites overlap heavily; together, 25 unique metabolites are significantly different between the pre- to post-TIPS samples. The only differentially abundant metabolite with an annotation in the pre- to post-PIV analysis was glycoursodeoxycholic acid (GUDCA), which decreases in the peripheral plasma immediately following the TIPS procedure (**Fig. 1f**). Meanwhile, 84 metabolites were differentially abundant post-PIV to BDc, with the majority mapping to annotated bile acids (**Fig. 1g**). Overall, these results demonstrate that TIPS placement is associated with changes in the abundance of bile acids in peripheral circulation.

### Post-TIPS HE severity is higher in participants with potentially less intrahepatic shunting

The surprisingly small difference observed between the pre- and post-HV metabolome suggested considerable intrahepatic shunting is already occurring in most participants prior to undergoing TIPS. The shunting is necessarily intrahepatic because the blood being sampled comes directly from the HV and thus would not include other portosystemic shunts such as esophageal varices or splenorenal shunts.

To explore how this could affect the future development of HE, we inspected metabolome changes based on the worst HE grade within a year after the TIPS procedure. Metabolome dissimilarities across participants post-TIPS show no significant differences for any of the pairwise HE grade comparisons (PERMANOVA all pairwise *P*>0.05; **Fig. 2a**), although there seems to be an increase in the spread of points with increasing severity. To ensure that this relationship between HE grade and variability was not due to reduction in portal hypertension after TIPS, we compared HE grade and changes in portal pressure and did not see a significant association (**Fig. 2b**).

**Fig. 2:**
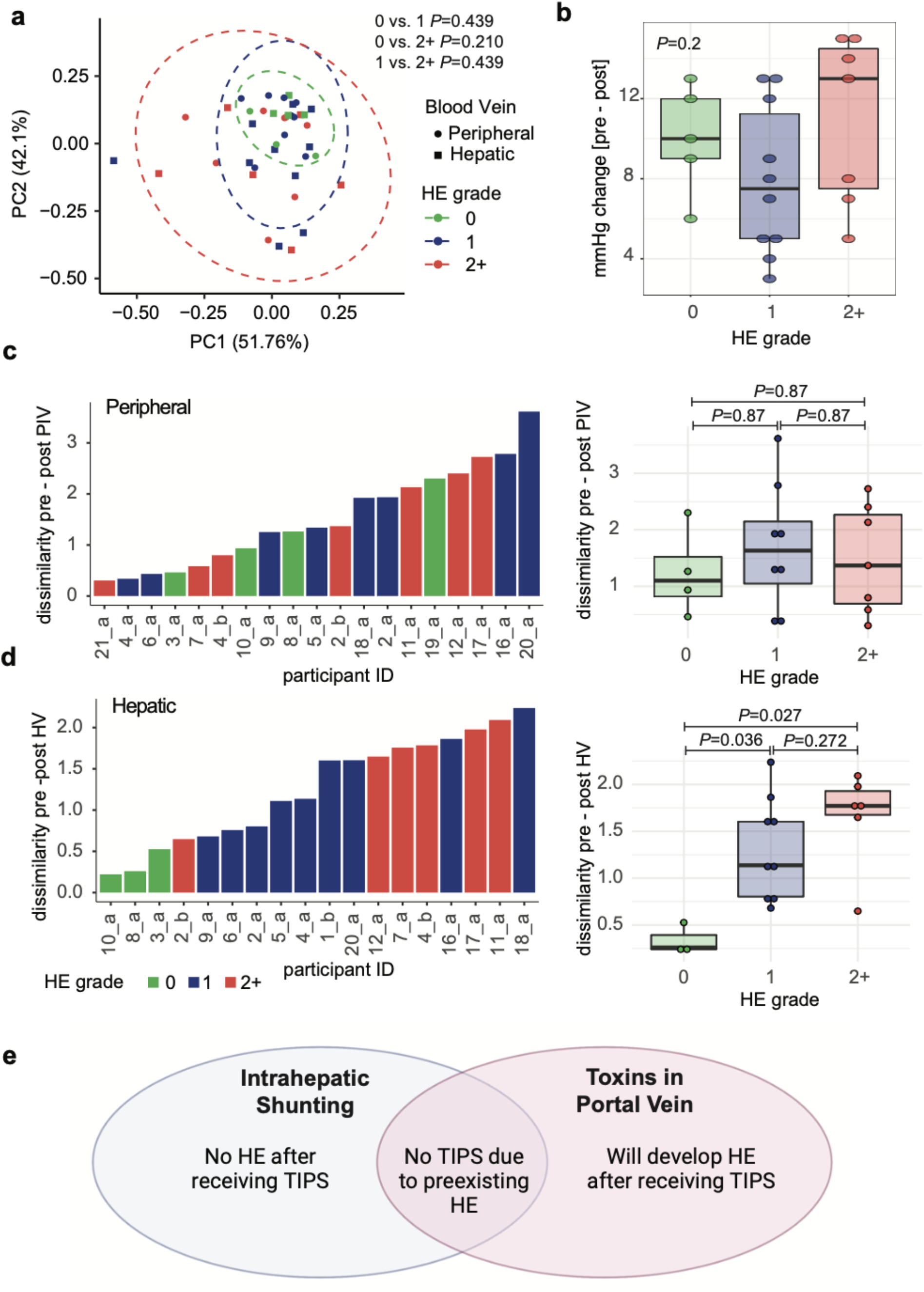
TIPS placement effect on metabolite patterns based on HE severity. **a**, RPCA plot showing metabolome dissimilarities across participants post-TIPS in PIV and HV samples stratified by HE grade. **b**, Portal pressure changes pre- to post-TIPS by HE grade. **c-d**, Comparison of the (**c**) PIV and (**d**) HV dissimilarity distances pre- to post-TIPS for each participant based on HE grade. Left: a ranked bar plot showing the (**c**) pre- to post-PIV or (**d**) pre- to post-HV dissimilarity distances for each participant. Right: boxplot showing the same data but grouped by participant HE grade. Dissimilarity is defined as the robust Aitchison β-diversity calculated with DEICODE. HE grade: 0=none (green); 1=mild (blue); 2+=severe (red). **e**, theoretical framework on the relationship between intrahepatic shunting, portal neurotoxins, and development of HE.

We then hypothesized that participants who develop more severe HE have less intrahepatic shunting at the time of their TIPS. To test this hypothesis, we compared the shift in metabolome within each participant before and after the TIPS procedure. In this analysis, those who already have significant intrahepatic shunting should have a low dissimilarity between their pre- and post-TIPS HV plasma metabolome, while those who have less shunting should have a higher dissimilarity. Within-participant dissimilarities between pre- and post-PIV metabolome did not show any difference based on HE grade (**Fig. 2c**). However, within-participant dissimilarities between pre- and post-HV metabolome are significantly different between participants with an HE grade of 0 versus participants with a grade of 1 or 2+ (Wilcoxon *P*=0.027 and *P*=0.036, respectively, **Fig. 2d**). In a retrospective power analysis we found that our sample sizes are sufficient to find a difference of 82% between HE grade 0 and the other groups (1-beta = 80%; alpha 5%). The fact that our actual differences between HE grade 0 and 1 is 391% and HE grade 0 and 2+ is 492% demonstrates that we have more than enough power in our sample size to detect differences in our groups. This change in the HV metabolome based on HE grade suggests that individuals with more dissimilarity in their metabolome after shunt placement, likely from a lack of pre-TIPS intrahepatic shunting, are at a higher risk of developing HE after the procedure (**Fig. 2e**).

### More severe HE outcomes are associated with decreased post-shunt levels of bile acids and glycerophosphocholine

Our analyses indicate that participants who developed HE had increased dissimilarity in their HV metabolome between their pre- and post-TIPS samples (**Fig. 2d**). To determine which metabolites are related to later development of HE, we analyzed how individual metabolites changed from baseline pre-TIPS levels (also called change from baseline and defined as log10(abundance_post_/abundance_pre_) for PIV or HV samples. This analysis revealed that the change in individual metabolite levels in participants who develop post-TIPS HE (grade 1 or 2+) is quite different from that of participants with no symptoms of HE (grade 0, **Fig. 3a-b**). Overall, participants with more severe HE (2+ grade) exhibit a different distribution of the change from baseline levels of their metabolites (**Fig. 3b**). This is observed for both hepatic and peripheral samples and characterized by an increased number of metabolites exhibiting lower change from baseline for participants who develop severe HE.

**Fig. 3:**
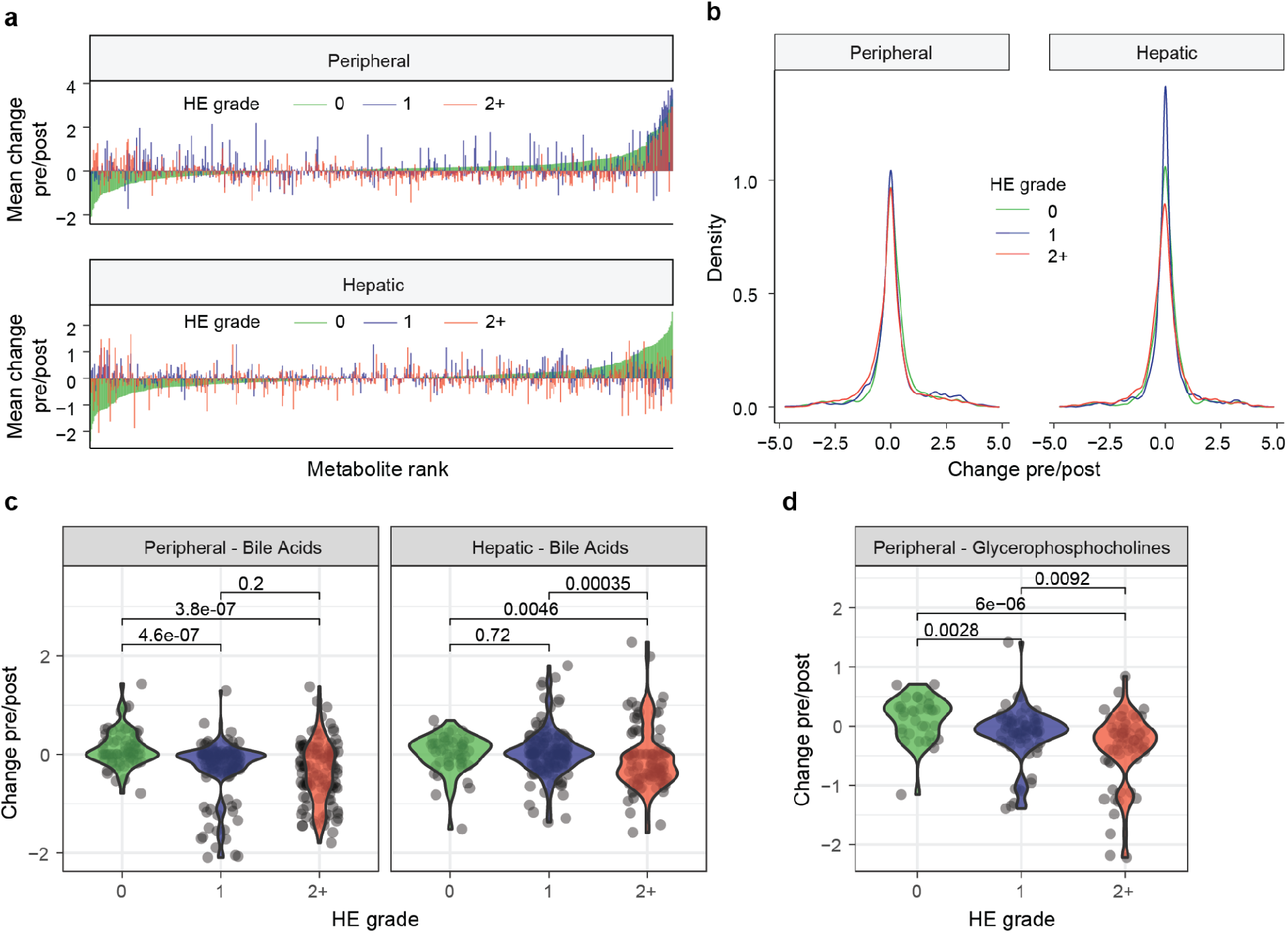
Change from baseline (pre/post) for individual metabolites. **a**, Mean change in pre/post abundance for each metabolite based on HE grade for peripheral or hepatic vein samples. Features are ordered in ascending order for HE grade “0” group. **b**, Density plot of the change pre/post in metabolite abundances of each participant based on HE grade. Kolmogorov–Smirnov test peripheral: 0 vs 2+, *P* = 1.3e-13; 0 vs 1, *P* = 1.0e-03; 1 vs 2+, P = 2.4e-09; hepatic: 0 vs 2+, *P* = 5.7e-08; 0 vs 1, *P* = 0.08; 1 vs 2+, *P* = 2.2e-16. **c-d**, Change pre/post TIPS for bile acids (peripheral and hepatic vein samples) (c) and glycerophosphocholines (peripheral samples) (d) between participants based on their HE grade (0, 1, or 2+). Change pre/post is defined as the log10(abundance_post_/abundance_pre_) for each participant’s metabolites. HE grade: 0 = none (green); 1 = mild (blue); 2+ = severe (red).

Out of 64 clusters (groups of related metabolites, **Supplementary Fig. S1**), two clusters with annotated compounds had significant differences based on HE severity: bile acids and glycerophosphocholines (**Fig. 3c-d and Supplementary Table 3**). Larger decreases from baseline in bile acids were observed for participants with more severe HE in peripheral and hepatic vein samples, while a similar trend for glycerophosphocholine was observed in peripheral plasma (**Figure 3c-d**). Changes from baseline levels of additional clusters are significantly different between HE grades but were not associated with HE severity (**Supplementary Table 3**).

### Low levels of post-shunt bile acids are associated with increased hepatic encephalopathy severity

Our results thus far demonstrate that bile acids exhibit differences from baseline levels that are related to development of HE. This suggests that these metabolites play a role in HE pathophysiology (**Fig. 3**), and their detection in PIV may help distinguish patients who may be at a high risk for post-TIPS HE. A closer examination of the bile acid metabolome showed that the abundances of three conjugated di- and tri-hydroxylated bile acids in the post-PIV are significantly correlated with HE grade. All three decrease with higher HE grade, suggesting that circulating levels of these bile acids post-shunt placement are inversely associated with HE severity (**Fig. 4a, Supplementary Table 4**).

**Fig. 4:**
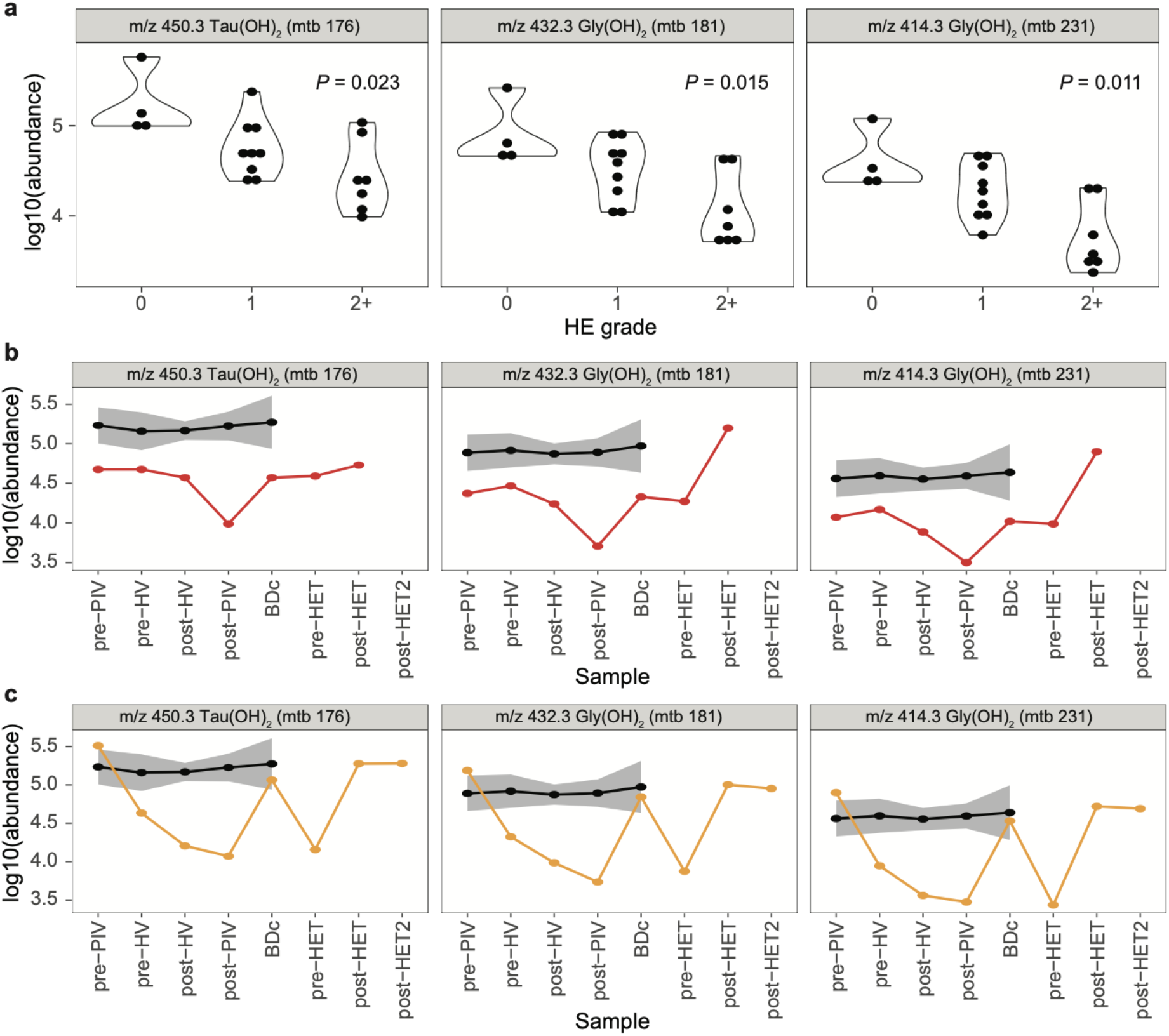
Levels of bile acids in plasma of TIPS participants based on HE grade. **a**, Bile acid levels and significant abundance differences in the post-TIPS peripheral blood based on HE grade. HE grade: 0=none; 1=mild; 2+=severe (Kruskal-Wallis test, FDR < 0.2). **b-c**, Longitudinal levels of bile acids in two participants readmitted with HE grade 2+ represented by red (b, participant 11_a) and orange (c, participant 12_a) compared to participants with HE grade 0 (black line, n=4; shaded areas represent SEM).

We next analyzed longitudinal bile acid levels for two participants (participants 11_a and 12_a) who were readmitted to the hospital within one year for HE treatment (HET) following a TIPS procedure. We investigated whether these three particular bile acid levels were also impacted at the time of readmission and treatment for HE (**Fig. 4b-c**). While these results are representative of only two participants, it is noteworthy that there is a drop in the PIV levels of these bile acids immediately after shunt placement in agreement with the results based on their later HE grade (**Fig. 4a-c**). Interestingly, upon readmission due to HE symptoms, bile acid levels are lower for participant 11_a, and HE treatment increases bile acid levels compared to pre-HET (**Fig. 4b**). An increase in these bile acids post-HET following readmission is also demonstrated for participant 12_a (**Fig. 4c**). The clinical course, demographics, and etiology for each readmitted participant revealed no overt predisposing clinical factors to explain the difference in the participants’ bile acid levels. Thus, these hydroxylated conjugated bile acids, which we suspect are secondary bile acids, may play a role in HE pathophysiology or predict which patients will develop worse HE.

### Abundance and mass shifts in glycerophosphocholines and bilirubins pre- to post-TIPS suggest potential hepatic biotransformations that prevent hepatic encephalopathy

The previous analyses demonstrated that TIPS placement disrupts the levels of specific metabolites in participants undergoing TIPS procedure (**Fig. 1c,e,f**), and that these changes are related to HE grade (**Fig. 3c-d** and **Fig. 4a**). We next sought to elucidate how shunt placement affects potential chemical transformations of related compounds and how they change over time. To this end, we used an approach called chemical proportionality (ChemProp), which takes advantage of longitudinal abundance data associated with FBMN (**Fig. 5a**).^37^ The ChemProp score allows us to prioritize the main changes in individual metabolite pairs based on high-scoring metabolite pairs, which in our case can point towards important functions that are being impacted with TIPS placement (**Fig. 5b**, ChemProp score). At the same time, by inspecting mass differences (**Fig. 5b**, delta m/z) between related metabolites, we highlight potential biotransformations that could be involved with these shifts in abundances (**Fig. 5b**, delta m/z annotation).

**Fig. 5:**
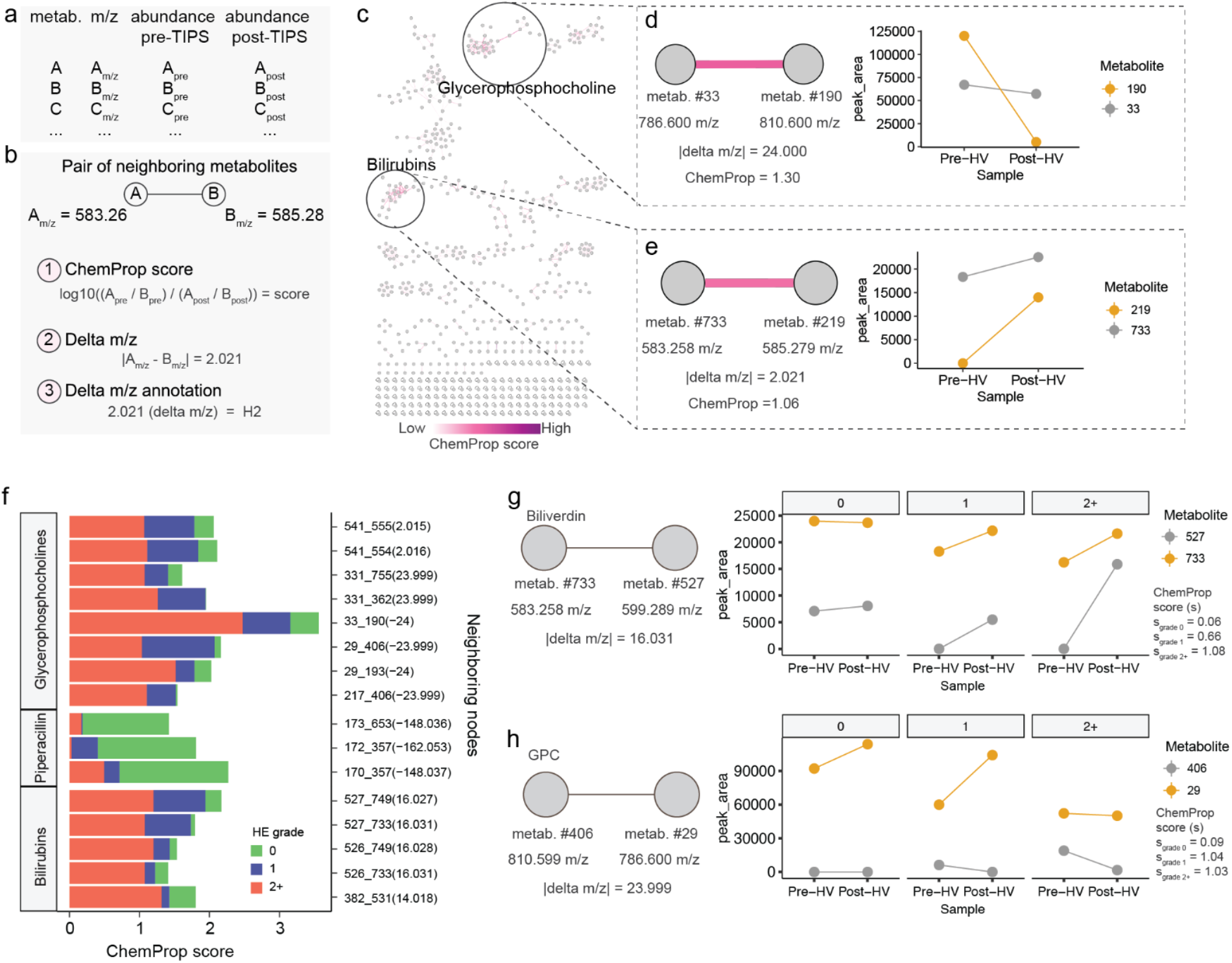
Chemical proportionality of neighboring metabolites pre- to post-TIPS placement. **a-b**, Schematic representation of the (a) quantification table from FBMN and (b) data that can be deduced from neighboring metabolites, including the ChemProp score, which is calculated as the log-ratio of two neighboring metabolites pre- and post-TIPS. **c**, Network representation of hepatic ChemProp scores highlighting high-scoring clusters. **d-e**, Specific examples of metabolite pairs within (d) glycerophosphocholines (GPC) and (e) bilirubins clusters that display high ChemProp scores. Metabolite ID and m/z are shown for metabolites, and associated delta m/z shown for metabolite pairs. **f**, ChemProp scores for pairs of neighboring metabolites that diverged the most on their scores based on HE grade. **g-h**, Examples of 2 metabolite pairs (within bilirubin and glycerophosphocholine subclasses) from panel f with divergent ChemProp scores based on HE grade. Median is shown for specific metabolites at each timepoint. HE grade: 0=none (green); 1=mild (blue); 2+=severe (red).

Upon calculating a ChemProp score for neighboring metabolites from hepatic or peripheral data, we further inspected some of the main changes pre- to post-shunt placement based on high-scoring pairs (**Fig. 5c and Supplementary Fig. S4**). The principal changes identified through ChemProp in the hepatic vein blood are related to metabolite pairs within glycerophosphocholine and bilirubin clusters, indicating the ratios between these metabolites fluctuate to a greater degree pre- to post-TIPS (**Fig. 5c**). For example, two diacylglycerophosphocholines with a mass change of 24.00 (C2) fluctuate to a high degree pre- to post-TIPS based on its ChemProp score (**Fig. 5d**). We also observe a high degree of fluctuation between two compounds within the bilirubin class, where one of the compounds matches biliverdin (**Fig. 5e**). The levels of these two compounds associated with a high ChemProp score and delta m/z of 2.021 suggests that biliverdin reduction is greatly impacted pre- to post-TIPS.

When taking HE grade into account, we additionally identified specific sets of metabolites fluctuating at different degrees pre- to post-TIPS in patients developing HE (**Fig. 5f**). For example, biliverdin oxidation could be implicated as contributing to post-TIPS HE, as the fluctuation between a metabolite matching biliverdin and another metabolite with a potential delta m/z 16.031 is high for patients with HE grade 2+ (**Fig. 5g**). Additionally, a mass shift of 23.99 (C2) between two diacylglycerophosphocholines is high in patients with severe HE (**Fig, 5h**). Interestingly, most bile acid ChemProp scores are low, suggesting that the TIPS procedure itself is not altering the ratios of bile acids pre- to post-TIPS (**Supplementary Fig. S4**).

## DISCUSSION

HE is a serious complication of liver disease, and TIPS placement can increase its risk. The etiology and treatment of HE have historically focused on ammonia homeostasis; however, ammonia levels are not universally predictive of the degree of HE and are not reliable in guiding treatment.^40,41^ In light of this incongruence, additional neurotoxins have been implicated, but no clinically applicable biomarker for HE severity has been validated. In this study, we used the latest methods in untargeted metabolomics with an expanded set of annotations to characterize metabolites implicated in changes post-TIPS that could lead to the development of HE. With further study, these compounds may be used as biomarkers and potential therapeutic targets for HE. Notably, the novel methods described herein are not limited to HE; they can be applied in virtually any disease process to identify involved metabolites, which can be investigated as biomarkers or targets for drug discovery.

Untargeted metabolomics allows us to compare metabolites within the peripheral and hepatic circulations before and after TIPS placement. We observed that the metabolome pre- to post-TIPS changes to a greater extent in the PIV compared to the HV (**Fig. 1b-c**), and the metabolites contributing the most to these changes are bile acids (**Fig. 1d-g, Supplementary Fig. 3**). Participants with the greatest degree of dissimilarity in the pre- to post-TIPS HV metabolome went on to develop HE (**Fig 2d)**. The higher degree of metabolite dissimilarity in HE grades 1 and 2+ implicates a functional and perhaps structural difference in participants who developed HE, namely, intrahepatic shunting. Our data strongly suggest that patients with a higher degree of intrahepatic shunting either have less portal toxins or are more acclimatized to the portal toxins prior to TIPS placement, and are therefore less likely to develop HE post-TIPS. Conversely, less intrahepatic shunting pre-TIPS may lead to an overwhelming level of portal toxins in the systemic circulation post-TIPs, with subsequent development of HE as a result (**Fig. 2e**). These findings imply that measurement of intrahepatic shunting pre-TIPS vs. post-TIPS could assist in risk-stratifying patients susceptible to developing high-grade HE months after TIPS.

The phenomenon of intrahepatic shunting is not widely acknowledged clinically, but has been described in literature. Specifically, the process is synonymous with pathologic hepatic angiogenesis, which is provoked by chronic liver disease.^6,42,43^ The microenvironment of inflammation, oxidative stress, shear stress, and hypoxia results in a synthetic milieu rich with angiogenic and endothelial growth factors.^44^ Vascular remodeling or “capillarization” occurs within hepatic sinusoids, resulting in altered hepatocyte blood flow. This can reduce the synthetic and detoxifying mechanisms within the previously functioning hepatocytes, as they are bypassed by the new capillaries.^45^ Methods of measuring intrahepatic shunts in patients with cirrhosis have been described, but have not been updated in decades. These involve imaging using radionucleotide-tagged galactose or ethanol injected into the hepatic veins and pulmonary arteries, angiography similar to that used for TIPS placement, and histologic stains for angiogenic factors after liver biopsy.^5,7,46^ Future work must be devoted to the development of readily-available, noninvasive clinical measurements of intrahepatic shunting within cirrhotic individuals before and after TIPS, as it may help determine which patients will be susceptible to the development of HE.

With the knowledge that intrahepatic shunting likely plays a role in post-TIPS HE and that the most significant metabolomic changes occur among bile acids pre- to post-TIPS, we explored the relationship between bile acids and HE. The change in bile acid abundances in the PIV and HV is greater in those who developed HE (**Fig. 3c**). This finding was confirmed upon examining bile acid changes as they relate to HE grade in the PIV, which showed three conjugated hydroxylated bile acids were inversely correlated with later development of HE (**Fig. 4a**). Patients who were hospitalized and treated for HE exacerbation had fluctuations in these specific bile acids that were consistent with their disease course (**Fig 4b-c**). Combined, our results suggest that increased risk of developing HE post-TIPS is related to the extent the bile acid abundance pool changes immediately post-TIPS, possibly in reference to these three bile acids. Closer inspection of these bile acids leads us to suspect that they are secondary conjugated glycine and taurine bile acids, with mtb 181 possibly corresponding to deoxycholic acid (DCA) or one of its isomers. These results suggest that bile acids play a protective role against the development of HE. Participants who did not develop post-TIPS HE did not demonstrate the decline in bile acids (**Fig. 3c & Fig. 4**), as they likely had intrahepatic shunting at the time of TIPS and had already acclimated to the portal toxins within systemic circulation. Those without intrahepatic shunting pre-TIPS are then exposed to the alterations in metabolites post-TIPS and are unable to further compensate for the portosystemic shunting.

Previous studies in animal models have shown that bile acids can give a therapeutic advantage. The neuronal oxidative damage induced by bilirubin can be inhibited by supplementation with the bile acid glycoursodeoxycholic acid (GUDCA).^47–51^ Indeed, the therapeutic, anti-inflammatory effects of GUDCA and TUDCA have been described in many disorders including insulin resistance, glucose intolerance, Barrett’s esophagus, retinal disease, and neurodegenerative diseases such as amyotrophic lateral sclerosis.^52–57^ GUDCA directly inhibited farnesoid X receptor, a ligand-mediated nuclear receptor whose activation is linked to HE and whose inhibition decreased HE symptoms in mice.^52,58^ To our knowledge, ours is the only study in humans which has linked bile acids as potentially protective against HE in humans.

Our analyses identified that specific bile acids decrease post-TIPS in participants with HE. With this knowledge, we turned to investigating chemical transformations in groups of metabolites that occur pre- to post-TIPS that could be effecting these changes. Through FBMN, we explored how mass changes in neighboring metabolite nodes implicate potential hepatic biochemical changes. Most of these changes occur among the bilirubin and glycerophosphocholine clusters (**Fig. 5d-g**). Specifically, our data show that oxidation-reduction reactions involving biliverdin occur to a greater degree in participants who developed high-grade HE. Modifications of glycerophosphocholine, a precursor to the important neurotransmitters acetylcholine and choline, have been previously described in patients with HE.^59–61^ However, the link between biliverdin oxidation-reduction and HE has not explicitly been studied, and warrants further exploration.

Biliverdin is involved in the catabolism of heme: heme-oxygenase 1 mediates the breakdown of heme into iron, carbon monoxide, and biliverdin; biliverdin is then reduced to bilirubin via biliverdin reductase A.^62,63^ Bilirubin is then bound to albumin for transport to the liver, where it is conjugated to glucuronic acid and excreted in bile.^64^ Bilirubin that is not conjugated remains bound to albumin in plasma, with a small amount of bilirubin remaining in plasma as free bilirubin. This free bilirubin is lipid-soluble, can cross the blood brain barrier, and can induce neuronal damage.^65,66^ While the link between biliverdin and HE has not been explicitly elucidated, previous studies have described a strong relationship between HE and free bilirubin.^67–69^ Interestingly, free bilirubin is rapidly solubilized by bile acids *in vitro*, perhaps explaining the potential protective effects of bile acids observed in our study In the context of bilirubin metabolism, our FBMN data suggest that an increase in the chemical reduction of biliverdin is involved in the development of post-TIPS HE (**Fig. 5g)**.^70^

At present, HE is managed via the enteric pharmaceuticals, rifaximin and lactulose.^71,72^ These drugs affect the composition of the gut microbiome, thus implicating microbial metabolites or products as potential agents that can influence the onset of HE– an observation supported by fecal microbiota transplant studies.^1,73^ The relationship between the bile acid pool and gut microbiome is well-established, but the role of the microbiome in HE is not thoroughly understood.^74–77^ Future areas of research should explore how the bile acid, glycerophosphocholine, and bilirubin metabolites in our study relate to the microbiome by performing fecal collections before and after TIPS procedure.

There are a few note-worthy limitations to our study. Our approach has limited coverage of the metabolic space, and not all metabolites are captured by the extraction and LC-MS/MS methods. Furthermore, the annotation of most compounds are based on the spectral libraries, many of which only allow for reliable subclass annotation as opposed to specific compounds (putative by MS/MS matching to reference library).^78^ We are also making an assumption that the toxin inducing HE is a metabolite, but it could be a protein or another signal coming from the gut or inflammatory system. In addition, this study would benefit from being supplemented by association analyses between the microbiome and metabolomics data to verify that our metabolite hits are of microbial origin. Our results should also be further verified in larger cohorts of patients, especially those who are being hospitalized and treated for HE to determine whether these bile acids are universally decreased in HE and are potential biomarkers for development of post-TIPS HE. Furthermore, since we show that intrahepatic shunting could be playing a protective role against HE, future studies should look into methods to measure the extent of intrahepatic shunting before TIPS. Bedside to bench experiments should further characterize the chemical structure of these bile acids, study their effects on neuroinflammation, and determine whether their supplementation could result in a treatment for HE.

In summary, this study used the innovative methodology of untargeted metabolomics to identify changes in metabolites in participants with cirrhosis undergoing TIPS. Relating these changes to HE suggested physiological differences between those that developed HE and those that did not, which led us to propose that intrahepatic shunting may be occurring in patients who develop HE post-TIPS. In two patients who were readmitted for HE exacerbations during our study period, we saw decreases in bile acids immediately post-TIPS, implying a protective mechanism for these metabolites. Furthermore, we demonstrated that glycerophosphocholines and bilirubins, important neuromodulating metabolites, fluctuate within our participants’ hepatic circulation post-TIPS. Ultimately, the findings described herein will assist in identifying potential biomarkers for HE, as well as targets for therapeutics in prevention and treatment of this highly morbid condition.

## Data Availability

All data produced are available online at https://gnps.ucsd.edu/

## ACKNOWLEDGMENTS

Initial funds for the studies were provided by a pilot and feasibility grant from UL1 TR001442. ACDM is supported by R01 HL148801-02S1. SF was supported by the PD Soros Foundation. AlZ is supported by NIH K08 DK113244 and the UF Foundation-Robert Duggan Fund. AmZ is supported by Kavli Institute for Brain and Mind at UC San Diego, AASLD Liver Scholar Award, VA Merit BLR&D Award I01 BX005707, and NIH K08 DK102902, R03 DK114536, R21 MH117780, R01 HL148801, R01 EB030134, and U01 CA265719. All UC San Diego authors receive institutional support from NIH P30 DK120515, P30 DK063491, P30 CA014195, P50 AA011999, and UL1 TR001442. The funders had no role in study design, data collection and interpretation, or the decision to submit the work for publication. The contents do not represent the views of the U.S. Department of Veterans Affairs or the United States Government

## AUTHOR CONTRIBUTIONS

Conceptualization - AmZ; Methodology - ACDM, AA, DP, AlZ, AmZ; Formal Analysis - SFR, ACDM; Investigation - ACDM, SFR, JMG; Resources - AlZ, AmZ, PCD; Data Curation - ADCM, SFR, JMG; Writing (Original Draft) - SFR, ACDM, AMC; Writing (Review & Editing) - JMG, DP, AA, AlZ, AmZ; Visualization - ACDM, SFR; Supervision - AlZ, AmZ; Project Administration - AlZ, AmZ; Funding Acquisition - AlZ, AmZ.

## METHODS

### Cohort and study design

This is a multi-center prospective cohort study approved by the IRBs of the University of California, San Diego, and University of Florida. Patients scheduled to undergo elective transjugular intrahepatic portosystemic shunt (TIPS) due to severe side effects related to portal hypertension were identified by participating interventional radiology physicians prior to the procedure. Patients undergoing TIPS were screened between August 2018 and February 2019. Inclusion criteria were met if patients were cirrhotic and undergoing elective TIPS, absence of HE at the time of enrollment, age >18, not pregnant, and willing and able to consent to the study. Patients were excluded if found to have non-cirrhotic portal hypertension, other potential causes of cognitive deficits, a previous liver transplant, or prescribed medications that could cause changes in the bile acid pool (e.g. ursodiol, sequestrants). Once study participants were selected for enrollment, informed consent was obtained by a study coordinator. On the day of the procedure, peripheral vein blood (PIV) was drawn from the patients pre-TIPS. During the procedure, the interventional radiologist collected hepatic vein (HV) blood just prior to and immediately after shunt placement, described as pre-TIPS and post-TIPS HV, respectively. Hepatic pressure in mmHg was recorded during TIPS as follows: gradient pre-TIPS was measured as the difference between the hepatic wedge pressure and the right atrium; gradient post-TIPS was measured as the difference between the portal vein pressure and the right atrium pressure. All participants received one 3.375g intravenous piperacillin-tazobactam dose intraoperatively. After the procedure but prior to admission to the post-anesthesia unit, the participants provided another peripheral vein blood sample (post-PIV). Finally, participants provided a fasting blood sample on the day of discharge with their morning labs (before discharge, BDc). Overall, the time difference between pre-PIV to post-PIV was 1-3 hours and post-PIV to BDc was 14-19 hours. The blood samples were centrifuged at 1,000g for 15 minutes at room temperature to obtain plasma and buffy coat, which were then aliquoted and stored at - 80 ?. Upon discharge, subjects were monitored for up to one year for readmission related to HE, and the worst HE grade (as defined by the West Haven criteria) during that period for each participant was noted.^79^ Additional peripheral blood samples were collected from participants readmitted during this time period.

After the data were collected, the hospital courses of the two participants who were readmitted during the study period were examined. Participant 11_a (**Fig. 3c**) was a 68-year-old woman with a history of cryptogenic cirrhosis with banded esophageal varices but with no prior history of HE. She underwent TIPS procedure due to progressively worsening ascites refractory to diuretic therapy and need for intermittent paracentesis. She was readmitted to the medical intensive care unit 9 days post-TIPS with obtundation requiring intubation for airway protection. Blood was drawn on the day of admission before initiation of hepatic encephalopathy treatment (pre-HET, **Fig. 3c**). Ammonia level on admission was 140 μmol/L. She was given lactulose and rifaximin for HE treatment, to which she responded well. Blood was collected for analysis on the day after treatment initiation (post-HET, **Fig. 3c**). She was discharged 9 days later. However, she was readmitted within 72 hours for altered mental status. Ammonia level at the time was 169 μmol/L, but she was less encephalopathic than prior admission and did not require intubation. Her encephalopathy resolved after continuous polyethylene glycol infusion via nasojejunal tube, but was again admitted 352 days post-TIPS for HE in the setting of urosepsis.

Participant 12_a (**Fig. 3b**) was a 62-year-old man with a history of alcoholic cirrhosis and esophageal varices, and with no history of HE. He underwent TIPS placement with simultaneous coil embolization of a left gastric varix. On post-TIPS day 30, he became acutely altered and was admitted to a nearby hospital, where ammonia level was 123 μmol/L. He was started on lactulose, but due to worsening encephalopathy, was transferred to the tertiary facility participating in the study. On admission to the participating facility, his ammonia level was 9 μmol/L, despite ongoing encephalopathy. Blood was collected prior to hepatic encephalopathy treatment (pre-HET, **Fig. 3b**). He underwent nasojejunal tube placement and was given polyethylene glycol continuously in addition to lactulose and rifaximin until his altered mentation resolved. Blood was again collected for analysis (post-HET, **Fig 3b**).

### Untargeted metabolomics by LC-MS/MS

For untargeted LC-MS/MS, 400 μl of pre-chilled extraction solvent (100% MeOH with 1.25 μM sulfamethazine) was added to 100 μl of plasma. Samples were briefly vortexed and then incubated at -20 °C for 20min for methanol extraction, after which they were centrifuged for 15 min at 20,000 x g. Supernatant was transferred to a pre-chilled 96-well deep well plate. Samples were dried using a centrifugal low pressure system for 8h. Dried extracts were stored in sealed plates at -80 °C until analysis. Samples were resuspended in methanol (MeOH) with 1.5 μM of sulfadimethoxine, vortexed, sonicated, transferred to a shallow 96-well plate, and diluted 2X. Untargeted metabolomics analysis was conducted using an ultrahigh-performance liquid chromatography system (UltiMate 3000; Thermo Fisher Scientific, Waltham, MA) coupled to a Maxis quadrupole time of flight (Q-TOF) mass spectrometer (Bruker Daltonics, Bremen, Germany) with a Kinetex C18 column (Phenomenex, Torrance, CA, USA). Data were collected as described in Gauglitz et al. 2020 in positive electrospray ionization mode.^80^ All solvents used were liquid chromatography-mass spectrometry [LC-MS]-grade (Fisher Scientific). Raw data were exported to open format .mzXML files using DataAnalysis (Bruker) and files were uploaded to the GNPS platform.

### Metabolomics data processing and analysis

Feature based molecular networking (FBMN) was performed on the GNPS platform using pre-processed MZmine2 files from LC-MS/MS experiments and is available under the following link:

https://gnps.ucsd.edu/ProteoSAFe/status.jsp?task=8068b7cb58f2465f97a15b3ce30d593c. In brief, MS/MS fragment ions +/- 17 Da of the precursor mz were removed and the top 6 fragment ions within a +/- 50 Da window were selected. Dataset and analysis files including MZmine2 parameters are available on MassIVE (dataset MSV000090443). Edges of the molecular network were created based on a filter of 0.65 cosine score minimum and more than 5 matched peaks. GNPS spectral libraries were used to match spectra in the network to get level 2 and level 3 annotations.^78^ Cytoscape was used to visualize networks. In addition, a chemical proportionality (ChemProp) approach was applied to the longitudinal data to calculate changes in abundance between neighboring nodes in order to identify changes in the abundances between every two neighboring nodes. The ChemProp score is calculated through the log_10_ transformed ratio of peak area proportions of two neighboring molecular network nodes between two sequential times points.^37^ This score reflects the level of the change in abundance pre-post TIPS for a given pair of neighboring metabolites, where a high score indicates greater changes, and the sign indicates directionality. Furthermore, inspecting the mass change associated with each pair and its ChemProp strength can indicate potential biological or chemical transformations within the network, which can be visualized on a dataset scale to prioritize modification patterns.^81^

### Metabolomics statistical analyses

To calculate metabolomic dissimilarities across participants, we used the robust aitchison β-diversity calculated using the DEICODE plugin in Qiime2 (--p-min-feature-count 10 and --p-min-sample-count 500) and plotted against robust principal component (RPCA) plots.^82,83^ We used a PERMANOVA test to find if there were significant differences between the metabolomes of subjects based on shunt placement. To find what metabolites were driving the dissimilarity between post-PiV and BDc in the RPCA, we used the tool Qurro to find what metabolite groups were most contributing to the directionality in the first principal component (PC1).^39^ Once we identified bile acids as being clustered to one side of PC1, we used Qurro to calculate the log ratio of bilirubins (a metabolite spread across PC1) to bile acids per sample. For each metabolite, we ran a paired Wilcoxon test on the log10(abundance) at Pre-PIV vs.Post-PIV, Post-PIV vs. BDc, and Pre-PIV vs. BDc, or Pre-HV vs. Post-HV using a FDR correction of 0.1. To determine statistically significant differences in the levels of bile acids based on the worst HE grade post-TIPS (0, 1, or 2+), we ran a Kruskal-Wallis test on the log10(abundance) of Post-PIV metabolites. Change from baseline or change pre/post was calculated as the log10(abundance_post_/abundance_pre_) for each participant’s metabolite. Statistical significance was determined for: a) all changes pre/post based on HE grade using a Kolmogorov–Smirnov test (0 vs 2+; 0 vs 1; 2+ vs 1); or b) changes pre/post for metabolites within each cluster using a Kruskal-Wallis test (0.2 FDR cutoff) followed by Wilcoxon test.

Custom scripts were used to plot within-participant metabolome dissimilarities pre to post shunt stratified by HE grade (code repository page will be made available for publication). All analyses were performed in R version 4.1.0.

## Data Availability

Dataset and analysis files are available on MassIVE under the MassIVE ID MSV000090443.

