## Supplemental Figure 1-4, Supplemental Table 1 for "Pre- and Post-Portosystemic Shunt Placement Metabolomics Reveal Molecular Signatures for the Development of Hepatic Encephalopathy"

********SUPPLEMENTARY MATERIAL********

Figure 1. Flowchart of patient screening, consent, admission
Figure 2. Feature-based molecular networking identifies metabolite features identified in plasma samples.
Figure 3. Qurro feature ranking.
Figure 4. Chemical proportionality pre- to post-TIPS.
Table 1. Demographic data of study participants

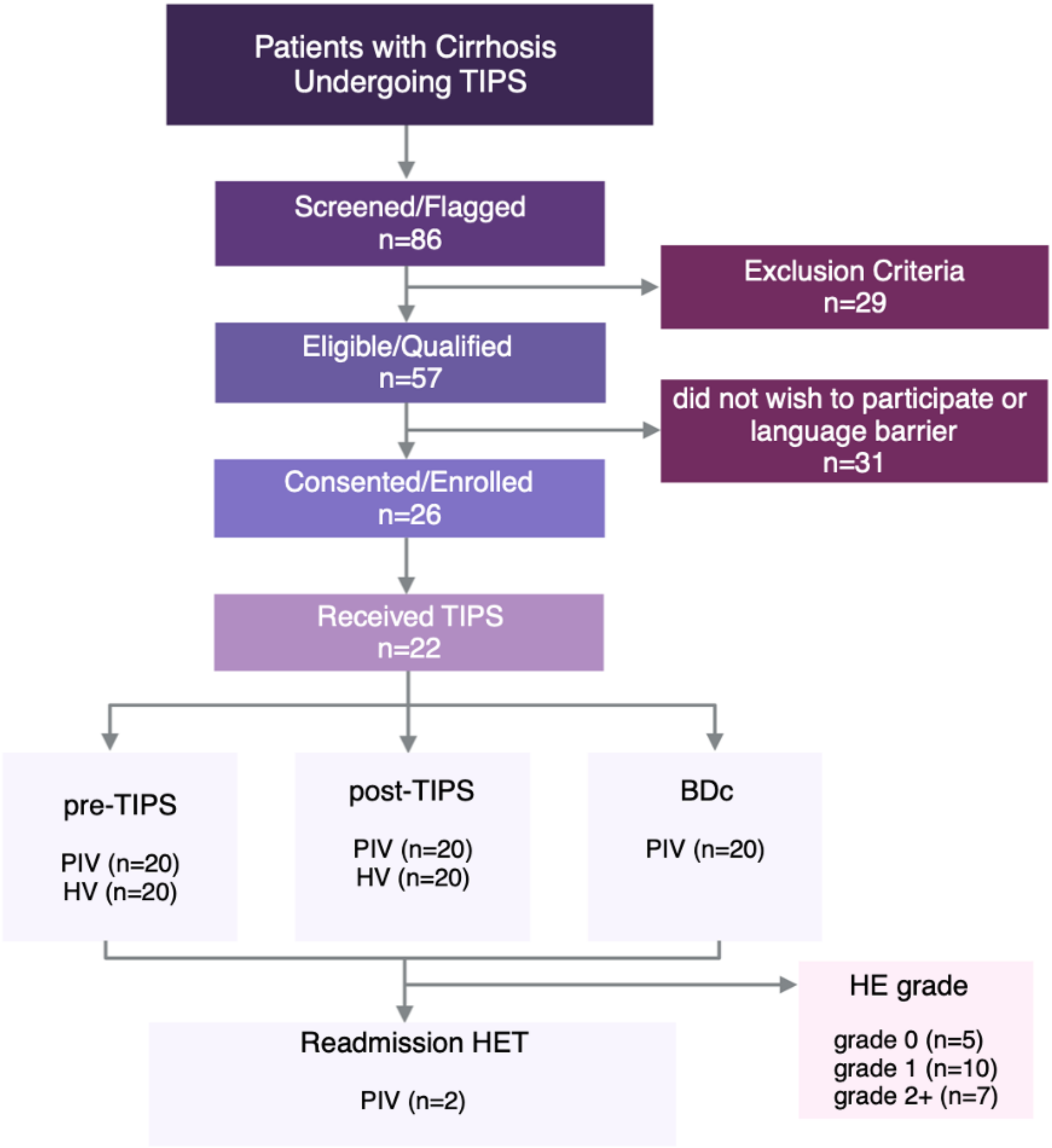


**Supplementary Figure 1.** Flowchart of patient screening, consent, admission. TIPS=transjugular intrahepatic portosystemic shunt, HE=hepatic encephalopathy, HET= hepatic encephalopathy treatment, PIV=peripheral vein, HV=hepatic vein. HE grade: 0=none; 1=mild; 2+=severe.

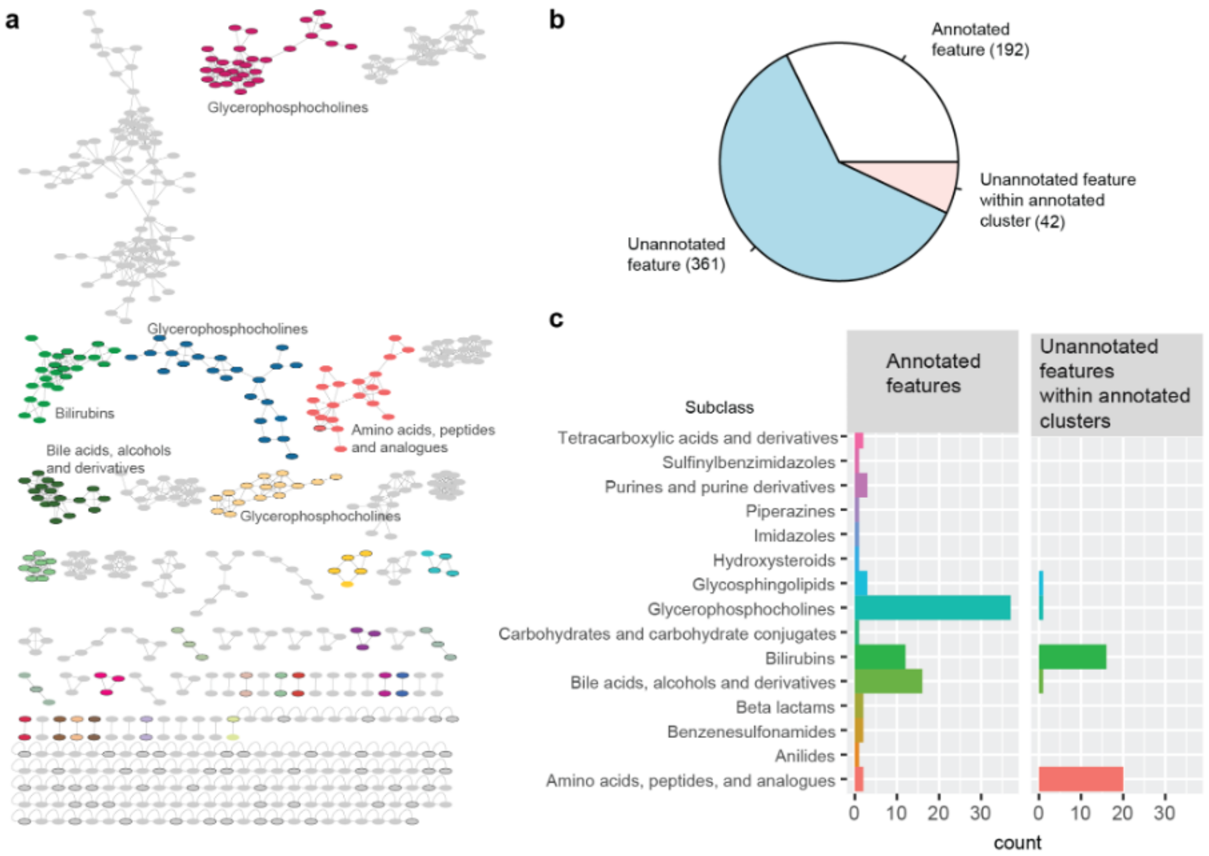


**Supplementary Figure 2.** Feature-based molecular networking identifies metabolite features identified in plasma samples. **a,** Molecular network from all samples. Each node represents a feature or metabolite. Lines connect neighboring nodes. Colors represent different clusters with at least two nodes and one annotated feature. Subclass of main annotated clusters are shown. **b,** Percentage of feature annotation: annotated features (n=192); features within annotated cluster (n=42); unannotated features (n=361). **c,** Representation of metabolite subclass classification based on GNPS annotation.

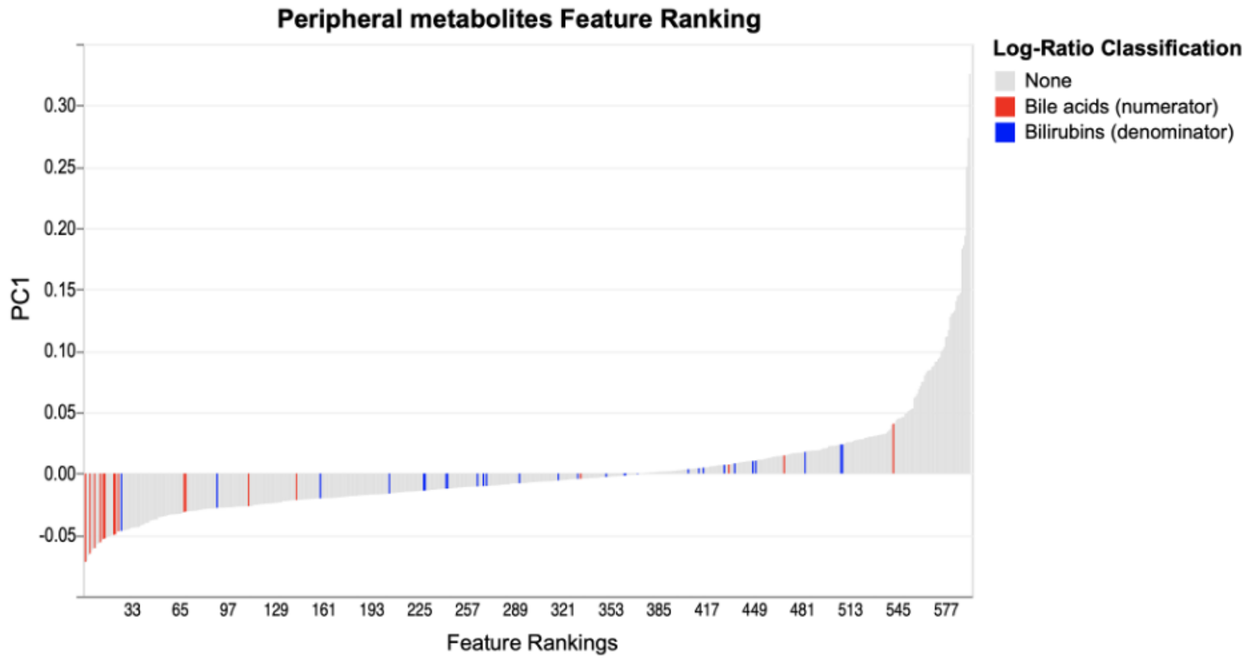


**Supplementary Figure 3.** Qurro feature ranking. The positioning of bile acids vs. bilirubins in the peripheral first principal component (PC1) for the DEICODE-calculated β-diversity was used to calculate the natural log ratio.

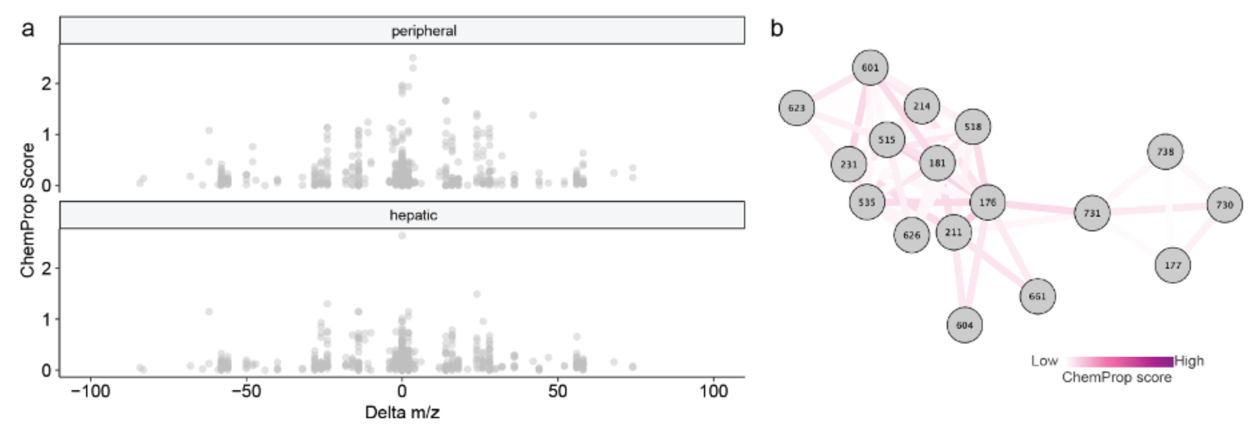


**Supplementary Figure 4.** Chemical proportionality pre- to post-TIPS. **a,** ChemProp score and associated Delta m/z for each pair of neighboring metabolites. **b,** Network representation of bile acids with associated IDs for individual metabolites and ChemProp scores for neighboring metabolite pairs.

**Supplemental Table 1.** Demographic data of study participants.

| Demographic Variable | participant total (N=22) |
|---|---|
| **Female (%)** | 8 (36%) |
| **Age (± SEM)** | 59.8 ± 2.4 |
| **Diagnosis* (%)** | |
| Alcohol | 10 (45%) |
| Cryptogenic | 3 (14%) |
| NASH | 8 (36%) |
| HCV | 2 (9%) |
| PBC | 1 (5%) |
| **MELD (± SEM)** | 12 ± 0.9 |
| **Indications for TIPS* (%)** | |
| Ascites | 20 (91%) |
| Esophageal Varices | 18 (82%) |
| Rectal Varices | 2 (9%) |
| **Post-TIPS HE (%)** | 17 (77%) |
| **Post-TIPS HE Grade (%)** | |
| 0 | 5 (23%) |
| 1 | 10 (45%) |
| 2+ | 7 (32%) |

*some patients had more than one diagnosis or indication for TIPS
NASH= nonalcoholic steatohepatitis; HCV= hepatitis C virus; PBC= primary biliary cirrhosis; MELD=Model for end stage liver disease; TIPS= transjugular intrahepatic portosystemic shunt; HE= hepatic encephalopathy. HE grade scored based on West Haven criteria.
